# Polygenic risk scores for the prediction of common cancers in East Asians: A population-based prospective cohort study

**DOI:** 10.1101/2022.09.12.22279874

**Authors:** Peh Joo Ho, Iain Bee Huat Tan, Dawn Qingqing Chong, Chiea Chuen Khor, Jian-Min Yuan, Woon-Puay Koh, Rajkumar Dorajoo, Jingmei Li

## Abstract

**Background:** To evaluate the utility of polygenic risk scores (PRS) in identifying high-risk individuals, different publicly available PRS for breast (n=65), prostate (n=26), colorectal (n=12) and lung cancers (n=7) were examined in a prospective study of 21,694 Chinese adults.

**Methods:** We constructed PRS using weights curated in the online PGS Catalog. PRS performance was evaluated by distribution, discrimination, predictive ability, and calibration. Hazard ratios (HR) and corresponding confidence intervals [CI] of the common cancers after 20 years of follow-up were estimated using Cox proportional hazard models for different levels of PRS.

**Results:** A total of 495 breast, 308 prostate, 332 female-colorectal, 409 male-colorectal, 181 female-lung and 381 male-lung incident cancers were identified. The area under receiver operating characteristic curve for the best performing site-specific PRS were 0.61 (PGS000004, breast), 0.66 (PGS00586, prostate), 0.58 (PGS000148, female-colorectal), 0.60 (PGS000734, male-colorectal) and 0.55 (PGS000740, female-lung), and 0.55 (PGS000392, male-lung), respectively. Compared to the middle quintile, individuals in the highest PRS quintile were 67% more likely to develop cancers of the breast, prostate, and colorectal. For lung cancer, the lowest PRS quintile was associated with 31-45% decreased risk compared to the middle quintile. In contrast, the hazard ratios observed for quintiles 4 (female-lung: 0.91 [0.58-1.44]; male-lung: 1.01 [0.74-1.38]) and 5 (female-lung: 1.00 [0.64-1.56]; male-lung: 1.07 [0.79-1.45]) were not significantly different from that for the middle quintile.

**Conclusions:** Site-specific PRSs can stratify the risk of developing breast, prostate, and colorectal cancers in this East Asian population. Appropriate correction factors may be required to improve calibration.

**Funding:** This work is supported by the National Research Foundation Singapore (NRF-NRFF2017-02), PRECISION Health Research, Singapore (PRECISE) and the Agency for Science, Technology and Research (A*STAR). WP Koh was supported by National Medical Research Council, Singapore (NMRC/CSA/0055/2013). CC Khor was supported by National Research Foundation Singapore (NRF-NRFI2018-01). Rajkumar Dorajoo received a grant from the Agency for Science, Technology and Research Career Development Award (A*STAR CDA - 202D8090), and from Ministry of Health Healthy Longevity Catalyst Award (HLCA20Jan-0022).

The Singapore Chinese Health Study was supported by grants from the National Medical Research Council, Singapore (NMRC/CIRG/1456/2016) and the U.S. National Institutes of Health [NIH] (R01 CA144034 and UM1 CA182876).

## INTRODUCTION

Polygenic risk scores (PRS) for a range of health traits and conditions have been developed in recent years. These scores, which are based on summary statistics from genome-wide association studies (GWAS), can be used to stratify people depending on their genetic risk of acquiring various diseases, to improve screening and preventative interventions, as well as patient care [1, 2]. Precision risk assessment may help develop tailored screening strategies targeting individuals at higher risk of disease of interest [3].

The contributions of heritable genetic factors are different for different cancers. Twin studies have highlighted statistically significant effects of heritable genetic risk factors for cancers of the prostate, colorectal, and breast [4]. The amount of phenotypic variance explained by the common genetic variants found by GWAS is also known to vary [5], suggesting that PRS derived from GWAS findings may perform to varying degrees for different cancers.

The area under receiver operating characteristic curve (AUC) is an important discrimination index for evaluating the performance of PRS. The greater the AUC, the better the discriminatory ability to separate cases from non-cases. A value of 0.5 suggests that the tool is performing no better than chance, while a value of 1 is obtained when cases and non-cases are perfectly separated. The range of reported AUC associated with published PRS ranged from 0.584 to 0.678 for breast cancer [6-12], 0.591 to 0.769 for prostate cancer [8, 10, 13], 0.609 to 0.708 for colorectal cancer [8, 10, 14, 15], and 0.52 to 0.846 for lung cancer [8, 10, 13, 16]. In a study by Jia et al looking at eight common cancers in the UK Biobank population-based cohort study (n=400,812 participants of European descent), the observed AUC ranged from 0.567 to 0.662 [10].

While prediction of individual cancer risks through PRS remains moderate, emerging data supports the use of PRS for population-based cancer risk stratification. In previous work, Ho et al examined the overlap of women identified to be at high risk of developing breast cancer based on family history for the disease, a non-genetic breast cancer risk prediction model, a breast cancer PRS, and carriership of rare pathogenic variants in established breast cancer predisposition genes [17]. The overlap of individuals found to be at elevated risk of developing breast cancer based on the genetic and non-genetic models was low. PRS was also found to be able to identify high-risk individuals among young women who were not yet eligible to attend mammography screening. The findings suggest that a genetic tool that is feasible to be deployed for population-based screening may complement current screening programs.

Disparities in the genetic risk of cancer among various ancestry populations are poorly understood. Ideally, selected genetic variants that make up PRS should be relevant to the population being screened. The development of training datasets of PRS are dominated by samples of European ancestry, resulting in ancestry bias and issues with transferability to other populations [2, 18]. The mismatch between the ancestries of the GWAS samples and the target populations for PRS application is a limiting factor [18]. In this study, we evaluated the utility of common PRS, curated in the Polygenic Score (PGS) Catalog, in predicting the risk of the commonly diagnosed cancers with high genetic predisposition (breast, prostate, colorectal, and lung) in a prospective cohort comprising 21,694 participants of East Asian descent in Singapore.

## METHODS

### Singapore Chinese Health Study (SCHS)

The Singapore Chinese Health Study (SCHS) is a population-based prospective cohort study of ethnic Chinese men and women recruited between April 1993 and December 1998 [19].

Participants were 45–74 years old at recruitment and were restricted to the two major dialect groups of Chinese adults in Singapore, who were the Hokkiens and the Cantonese that had originated from Fujian and Guangdong provinces in Southern China, respectively. All our study participants were residents of government housing flats, which were built to accommodate approximately 86% of the resident population in Singapore during the enrolment period. A total of 63 257 individuals (35,298 women and 27,959 men) provided written informed consent [19]. The study was approved by the Institutional Review Boards of the National University of Singapore, University of Pittsburgh, and the Agency for Science, Technology and Research (A*STAR, reference number 2022-042). Written, informed consent was obtained from all study participants.

#### Baseline

An in-person baseline interview was performed at recruitment to collect data on diet using a validated 165-item food frequency questionnaire, smoking, alcohol, physical activity, medical history, and menstrual and reproductive history from women.

#### Selection of common cancers

In Singapore, between 2015 and 2019, colorectal cancer, the most prevalent cancer in men, accounted for nearly 17% of cancer diagnoses, while breast cancer, the most common cancer in women, accounted for about three out of ten cancer diagnoses (Singapore Cancer Registry Annual Report 2018). During this time, cancers of the breast, prostate, colorectal, and lung accounted for approximately half of the total cancer diagnoses. These four most common cancers were selected for inclusion in this study.

A unique National Registration Identity Card (NRIC) number for every Singaporean enables the compilation and linkage of data from national register data to the same individual [20]. Identification of incident cases of cancer was accomplished by record linkage of all surviving cohort participants with the database of the nationwide Singapore Cancer Registry [20]. Cancers that developed among SCHS participants were identified using International Classification of Diseases (ICD) codes ICD-O-3 (breast: C50, prostate: C61, colorectal: C18, C19 and C20, lung: C34).

#### Follow-up

Death date was obtained by record linkage with the database Birth and Death Registry of Singapore [20]. To date, only 47 (<1%) of the entire cohort participants were known to be lost to follow-up due to migration out of Singapore, suggesting that the ascertainment of cancer and death incidences among the cohort participants was virtually complete.

### Genotyping and imputation

Between 1999 and 2004, a total of 28,346 subjects contributed blood samples. A total of 25,273 SCHS participants were genotyped between the years 2017 to 2018 with the Illumina Infinium Global Screening Array (GSA) v1.0 and v2.0 [21].

Details on the sample quality control (QC) processes are previously described [21]. Briefly, samples with a call rate of 95% or below (n=176) or heterozygosity extremes (>3 standard deviation, n=236) were removed. Identity-by-state measurements were performed by pairwise comparisons of samples to detect related samples (first and second degree). One sample from each identified pair with the lower call rate was eliminated from further analysis (n=2,746). To identify any ethnic outliers, principal component analysis (PCA) was used in conjunction with 1000 Genomes Project reference populations and within the SCHS samples, which resulted in the further removal of 287 samples. Of the 21,828 samples that passed genotyping quality control, 134 participants who were diagnosed with cancer before recruitment or had missing cancer outcomes and were excluded from the study, resulting in a final analytical dataset of 21,694 (**Supplementary Figure 1**).

Alleles for all SNPs were coded to the forward strand and mapped to hg19. SNP quality control steps included the exclusion of sex-linked and mitochondrial variants, gross Hardy–Weinberg equilibrium (HWE) outliers (P < 1 × 10^−6^), monomorphic SNPs or those with a minor allele frequency (MAF) < 1.0%, and SNPs with low call-rates (<95.0%). We imputed for additional autosomal SNPs using IMPUTE v2 [22] and with a two reference panel imputation approach by including 1) the cosmopolitan 1000 Genomes reference panels (Phase 3, representing 2,504 samples) and 2) an Asian panel comprising 4,810 Singaporeans (2,780 Chinese, 903 Malays, 1127 Indians) [21]. SNPs with imputation quality score INFO < 0.8, MAF < 1.0%, or HWE P < 1 × 10^−6^, as well as non-biallelic SNPs were excluded.

### Polygenic risk scores (PRS)

Published polygenic risk scores (PRS) were retrieved from The Polygenic Score (PGS) Catalog, an open database of polygenic scores (retrieved on Feb 26, 2022) (**Additional file 1 - Supplementary Table 1**) [23]. Of the 2,166 PRS available in the resource, 1,706 PRS comprising less than 100,000 predictors were downloaded. A total of 65, 26, 12, and 7 PRS were available for breast, prostate, colorectal, and lung cancers, respectively. **Additional file 1 - Supplementary Table 2** shows the number of individual variants comprising each PRS and proportion of variants missing in the SCHS cohort. Individual PRS were calculated using the allelic scoring (–score sum) functions with default parameters in PLINK (v1.90b5.2) [24].

### PRS distribution

Two-sided, two-sample t-tests with a type I error of 0.05 were used to examine whether there was a difference in the distribution of standardised PRS (subtraction of mean value followed by the division by the standard deviation) between site-specific cancer cases and non-cancer controls.

### PRS discrimination

Discrimination was quantified by the area under the receiver operating characteristic (ROC) curve (AUC), using logistic regression models, and their corresponding 95% CI. An AUC of 0.9–1.0 is considered excellent, 0.8–0.9 very good, 0.7–0.8 good, 0.6–0.7 sufficient, and 0.5–0.6 insufficient [25].

### Associations between PRS and risk of developing cancers

Subjects were classified into PRS percentile groups. Person-years of follow-up were calculated for each subject from the date of enrolment to the date of cancer diagnosis, death, or December 31, 2015 (the date of linkage with the Singapore Cancer Registry), whichever came first. Follow-up time was censored at 20 years after recruitment. The associations between PRS quintiles (where individuals ranked by PRS were categorised into quintiles, using the middle quintile [40 to 60%] as reference to reflect the average risk of the population) and the incidence of site-specific cancers were investigated using Cox proportional hazards modelling to estimate hazard ratios (HR) and corresponding 95% confidence intervals (CI), using time since recruitment as the time scale, and adjusted for age at recruitment. Tests for trends were conducted using two-sided Wald tests with a type I error of 0.05. Assumptions for proportional hazards were checked using the cox.zph() function in the “survival” package in R.

HR and corresponding 95% CI were also estimated for every standard deviation (SD) increase in PRS. Variables adjusted in the models included age at recruitment, dialect group (Hokkien or Cantonese), highest level of education (no formal education, primary school, or secondary or higher), body mass index (continuous, kg/m^2^), cigarette smoking (non-smoker, ex-smoker, current smoker), alcohol consumption (never, weekly, daily), moderate physical activity (none, 1-3h/week, ≥3h/week), vigorous work/strenuous physical activity at least once a week (no or yes), and familial history of cancer (no or yes).

To estimate the HR for each individual, we applied the *predict()* function with option *type=“risk”* to the Cox model with PRS (standardised to mean 0 and variance 1) and age at recruitment. The proportion of study participants in the cohort with a given relative risk of each site-specific cancer (HR_per SD increase in PRS_ = 1.5, 2.0, 2.5, and 3.0), and the percentage of at-risk individuals (based on the respective HR cut-offs) that develop cancer in all site-specific cancers were estimated.

### PRS predictive ability

The five-year absolute risks of developing breast, prostate, colorectal, and lung cancers were computed for PRS groups of increasing five percentiles over the follow-up period. Incidence (between 2013 to 2017) and mortality (the year 2016) statistics in Singapore (reported in [26] and [27], respectively) were used for the absolute risk estimations.

### PRS calibration

Calibration was studied by comparing the expected proportion of cases in the five years after recruitment to the observed proportion of cases that occurred in that five years, within each decile of PRS. Linear regression of the ten points (pairs of expected and observed proportion) was used to study the overall calibration. A curve close to the diagonal indicates that predicted cancer risks correspond well to observed proportions. A slope above 1 implies that the model underestimates the absolute risk. Conversely, a slope below 1 implies that the model overestimates the absolute risk.

## RESULTS

### Characteristics of the study population

**Table 1** shows the characteristics of the 21,694 participants who were cancer-free at recruitment. The median follow-up time for the cohort was 20 years (IQR: 18 to 22). As of December 2015, 495 women developed breast cancer, 308 men developed prostate, 774 (332 women and 409 men) colorectal cancer, and 562 (181 women and 381) lung cancer. The median age at recruitment was 54 years (interquartile range [IQR]: 49 to 61). The median age at diagnosis was 65 years (IQR: 59-70) for female breast cancers, 72 years (IQR: 67 to 77) for prostate cancers, 71 years (IQR: 65 to 76) for male colorectal cancers, 71 years (IQR: 64 to 78) for female colorectal cancers, 74 years (IQR: 68 to 78) for male lung cancers and 74 years (IQR: 66 to 79) for female lung cancers. Sixteen percent of the cohort (n=3,501) reported positive first-degree family history of any cancer at baseline interview.

**Table 1.** Demographics of our study population by gender and cancer site. Demographics variables were collected using structured questionnaire at recruitment. Family history for lung cancer was not available. Information on cancer occurrence (number of cancer and age at cancer occurrence) was obtained through linkage with the Singapore Cancer Registry in December 2015. Follow-up time was calculated from age at recruitment. IQR: Interquartile range.

|  | Entire cohort |  |  | Individuals who developed cancer |  |  |  |  |  |
| --- | --- | --- | --- | --- | --- | --- | --- | --- | --- |
|  | All | Female | Male | Breast | Prostate | Colorectal |  | Lung |  |
|  |  |  |  | Female | Male | Female | Male | Female | Male |
| *n* | 21694 | 12084 | 9610 | 495 | 308 | 332 | 409 | 181 | 381 |
| Age at recruitment in years, median (IQR) | 54 (49–61) | 54 (48–60) | 55 (49–62) | 53 (48–59) | 59 (54–64) | 58 (52–64) | 59 (52–65) | 59 (55–64) | 60 (55–64) |
| Number of cancers developed |  |  |  |  |  |  |  |  |  |
| 0 (did not develop cancer) | 19633 (90) | 11096 (92) | 8537 (89) | - | - | - | - | - | - |
| 1 | 2013 (9) | 968 (8) | 1045 (11) | 476 (96) | 293 (95) | 317 (95) | 387 (95) | 175 (97) | 362 (95) |
| 2 | 48 (0) | 20 (0) | 28 (0) | 19 (4) | 15 (5) | 15 (5) | 22 (5) | 6 (3) | 19 (5) |
| Age at diagnosis among individuals who develop cancer(s) (earliest age for those with multiple cancers) in years, median (IQR) | 70 (64–77) | 68 (62–76) | 72 (67–77) | 65 (59–70) | 72 (67–77) | 71 (64–78) | 71 (65–6) | 74 (66–79) | 74 (68–78) |
| Length of follow-up (longest follow-up for those with multiple cancers) in years, median (IQR) | 20 (18–22) | 20 (18–22) | 19 (17–21) | 11 (6–16) | 13 (9–17) | 13 (8–17) | 11 (7–16) | 14 (9–17) | 14 (10–17) |
| Dialect group (%) |  |  |  |  |  |  |  |  |  |
| Hokkien | 10663 (49) | 6132 (51) | 4531 (47) | 260 (53) | 153 (50) | 185 (56) | 164 (40) | 95 (52) | 162 (43) |
| Cantonese | 11031 (51) | 5952 (49) | 5079 (53) | 235 (47) | 155 (50) | 147 (44) | 245 (60) | 86 (48) | 219 (57) |
| Highest education (%) |  |  |  |  |  |  |  |  |  |
| No | 4629 (21) | 3878 (32) | 751 (8) | 128 (26) | 20 (6) | 123 (37) | 46 (11) | 85 (47) | 57 (15) |
| Primary level | 9760 (45) | 5082 (42) | 4678 (49) | 206 (42) | 146 (47) | 138 (42) | 232 (57) | 62 (34) | 228 (60) |
| Secondary or above | 7305 (34) | 3124 (26) | 4181 (44) | 161 (33) | 142 (46) | 71 (21) | 131 (32) | 34 (19) | 96 (25) |
| Body mass index in kg/m<sup>2</sup>, median (IQR) | 23 (21–25) | 23 (21–25) | 23 (21–25) | 23 (21–25) | 23 (21–25) | 23 (21–24) | 23 (21–25) | 23 (20–24) | 23 (20–24) |
| Smoking status (%) |  |  |  |  |  |  |  |  |  |
| Never | 15553 (72) | 11235 (93) | 4318 (45) | 472 (95) | 166 (54) | 296 (89) | 153 (37) | 129 (71) | 63 (17) |
| Ex-smoker | 2374 (11) | 261 (2) | 2113 (22) | 8 (2) | 66 (21) | 14 (4) | 108 (26) | 9 (5) | 74 (19) |
| Current smoker | 3767 (17) | 588 (5) | 3179 (33) | 15 (3) | 76 (25) | 22 (7) | 148 (36) | 43 (24) | 244 (64) |
| Number of cigarettes smoked (%) |  |  |  |  |  |  |  |  |  |
| Does not smoke | 15553 (72) | 11235 (93) | 4318 (45) | 472 (95) | 166 (54) | 296 (89) | 153 (37) | 129 (71) | 63 (17) |
| <12 | 2408 (11) | 581 (5) | 1827 (19) | 14 (3) | 54 (18) | 26 (8) | 85 (21) | 36 (20) | 81 (21) |
| 13–22 | 2344 (11) | 206 (2) | 2138 (22) | 6 (1) | 53 (17) | 9 (3) | 108 (26) | 15 (8) | 135 (35) |
| >=23 | 1389 (6) | 62 (1) | 1327 (14) | 3 (1) | 35 (11) | 1 (0) | 63 (15) | 1 (1) | 102 (27) |
| Alcohol consumption (%) |  |  |  |  |  |  |  |  |  |
| Never/ occasionally | 19079 (88) | 11506 (95) | 7573 (79) | 470 (95) | 253 (82) | 315 (95) | 303 (74) | 174 (96) | 296 (78) |
| Weekly | 1885 (9) | 437 (4) | 1448 (15) | 20 (4) | 44 (14) | 10 (3) | 66 (16) | 5 (3) | 49 (13) |
| Daily | 730 (3) | 141 (1) | 589 (6) | 5 (1) | 11 (4) | 7 (2) | 40 (10) | 2 (1) | 36 (9) |
| Moderate physical activity (%) |  |  |  |  |  |  |  |  |  |
| No | 16584 (76) | 9446 (78) | 7138 (74) | 380 (77) | 208 (68) | 269 (81) | 295 (72) | 143 (79) | 294 (77) |
| 1 to 3 hours/week | 3274 (15) | 1679 (14) | 1595 (17) | 69 (14) | 62 (20) | 43 (13) | 68 (17) | 23 (13) | 53 (14) |
| >= 3 hours/week | 1836 (8) | 959 (8) | 877 (9) | 46 (9) | 38 (12) | 20 (6) | 46 (11) | 15 (8) | 34 (9) |
| Vigorous physical activity/ strenuous sports at least once a week (%) |  |  |  |  |  |  |  |  |  |
| No | 18467 (85) | 11221 (93) | 7246 (75) | 452 (91) | 239 (78) | 311 (94) | 342 (84) | 175 (97) | 314 (82) |
| Yes | 3227 (15) | 863 (7) | 2364 (25) | 43 (9) | 69 (22) | 21 (6) | 67 (16) | 6 (3) | 67 (18) |
| Family history of any cancer in first degree relatives (%) |  |  |  |  |  |  |  |  |  |
| No | 18193 (84) | 10141 (84) | 8052 (84) | 404 (82) | 236 (77) | 281 (85) | 336 (82) | 165 (91) | 333 (87) |
| Yes | 3501 (16) | 1943 (16) | 1558 (16) | 91 (18) | 72 (23) | 51 (15) | 73 (18) | 16 (9) | 48 (13) |

Overall, eight in ten participants (79%) reported an education level of primary school and above. However, the proportion of females who did not receive an education (32%) was four times higher compared to males (8%). Median BMI was 23 kg/m^2^ in the overall cohort, as well as in sex specific and site-specific subgroups. There were more non-smokers among females (93%) compared to males (45%). Alcohol consumption was low among the participants, with 88% of the cohort reported never or occasional drinking (79% male, 95% female). Three in four participants reported regular engagement in moderate physical activity; 85% of the participants reported no participation in higher levels of physical activity.

### Lack of Asian representation in PRS development

Among PRS for breast (n=65), prostate (n=26), colorectal (n=12) and lung cancers (n=7) examined, the reported source of variant associations or GWAS used to build PRS were from predominantly European ancestry populations (**Additional file 1 - Supplementary Table 2**). Only one PRS for breast cancer (PGS001778) and two PRS for colorectal cancer (PGS000802 and PGS000734) were based on GWAS that included some non-European participants. For PRS development training, all but two PRS were based on samples of non-European ancestry (PGS000733 for prostate cancer and PGS000802 for colorectal cancer). No significant association (P>0.05) was found between number of variants included in the various PRS evaluated for each cancer and discriminatory ability (**Additional file 1 - Supplementary Table 3**).

### PRS distribution

**Figure 1** depicts the A) distribution, B) discrimination, C) predictive ability, and D) calibration of the best-performing PRS (based on AUC) (**Additional file 1 - Supplementary table 3**) for the four cancers studied: breast (PGS000004), prostate (PGS00586), colorectal (female: PGS000148; male: PGS000734), and lung (female: PGS000740; male: PGS000392). All PRS were normally distributed, with a right shift observed in the distribution curves for cancer cases (**Figure 1A**). The mean value of each site-specific cancer PRS was significantly higher in cancer patients compared to controls (P_t-test_<0.00273).

**Figure 1.**
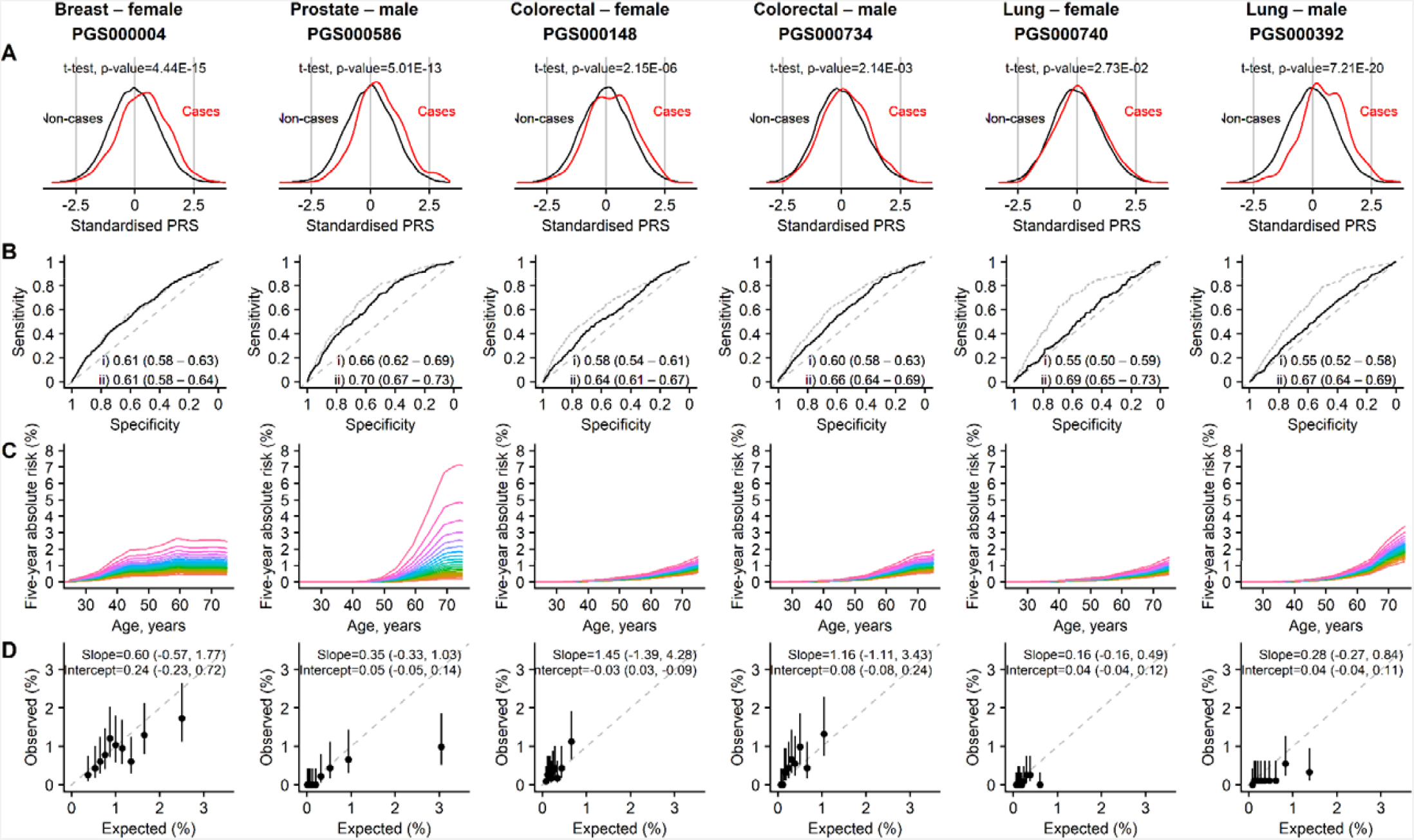
Site-specific polygenic risk scores (PRS) performance assessment. A) Distribution, B) discrimination, C) predictive ability and D) calibration for each of the four common cancers studied. Two-sided, two-sample t-tests with a type I error of 0.05 were used to examine whether there was a difference in the distribution of standardised PRS (subtraction of mean value followed by the division by the standard deviation) between site-specific cancer cases and non-cancer controls (A). The PRS showcased are the best-performing scores based on Area Under the Receiver Operator Characteristic Curve (AUC) values in the female and male populations, i) unadjusted [solid line], and ii) adjusted for age at recruitment [dashed line] (B). Each colored line in the plots for predictive ability denotes a five percentile increase in the standardised PRS score in (C). Calibration calculated based on five-year absolute risk by PRS deciles in (D). A prediction tool is considered more accurate when the AUC is larger. An AUC of 0.9–1.0 is considered excellent, 0.8–0.9 very good, 0.7–0.8 good, 0.6–0.7 sufficient, 0.5–0.6 bad, and less than 0.5 considered not useful (PMID: 27683318).

### Associations between PRS and risk of developing cancers

During the follow-up period of 20 years, the risk of acquiring breast, colorectal, or lung cancer increased significantly with higher PRS after adjusting for age at recruitment. Compared to the first PRS quintile, individuals in the highest quintile were more likely to develop the four cancers studied. The highest hazard ratio observed was for prostate cancer (4.72 [95%CI: 3.04 – 7.34]) and lowest for male lung cancer (1.54 [1.10 – 2.16]), adjusted for age at recruitment (**Additional file 1 - Supplementary Table 4**). Significant trends were found for the associations between PRS quintiles and site-specific cancers (P-trend ranges from 7.30 × 10^−17^ for prostate cancer to 0.029 for female lung cancer, **Additional file 1 - Supplementary Table 4**).

Compared to the middle PRS quintile, individuals in the highest PRS quintile were more than 67% more likely to develop cancers of the breast, prostate, and colorectal (**Table 2**). Individuals in the lowest PRS quintile were associated with a 30-65% reduction in risk of developing these cancers. For lung cancer, the lowest PRS quintile was associated with 31-45% decreased risk compared to the middle quintile. However, the hazard ratios observed for quintiles 4 (female: 0.91 [0.58 to 1.44]; male: 1.01 [0.74 to 1.38]) and 5 (female: 1.00 [0.64 to 1.56]; male: 1.07 [0.79 to 1.45]) were not significantly different when compared to the middle quintile.

**Table 2.** Hazard ratios (HR) and corresponding 95% confidence intervals (CI) associated with polygenic risk score quintiles (Q) compared to the population median, using the Cox proportional hazards model and censored at 20 years after recruitment. All models were adjusted for age at recruitment.

| Cancer site - gender | Q1 | Q2 | Q3 | Q4 | Q5 |
| --- | --- | --- | --- | --- | --- |
| Breast – Female |  |  |  |  |  |
| Number of cases | 54 | 76 | 86 | 103 | 147 |
| HR (95%CI) | 0.60 (0.43 – 0.84) | 0.84 (0.62 – 1.14) | 1.00 (Referent) | 1.19 (0.89 – 1.58) | 1.67 (1.28 – 2.17) |
| Prostate – Male |  |  |  |  |  |
| Number of cases | 24 | 27 | 68 | 59 | 111 |
| HR (95%CI) | 0.35 (0.22 – 0.57) | 0.42 (0.27 – 0.65) | 1.00 (Referent) | 0.89 (0.63 – 1.26) | 1.67 (1.24 – 2.26) |
| Colorectal – Female |  |  |  |  |  |
| Number of cases | 37 | 63 | 51 | 74 | 85 |
| HR (95%CI) | 0.70 (0.46 – 1.07) | 1.25 (0.87 – 1.81) | 1.00 (Referent) | 1.48 (1.04 – 2.12) | 1.69 (1.19 – 2.39) |
| Colorectal – Male |  |  |  |  |  |
| Number of cases | 36 | 70 | 71 | 87 | 114 |
| HR (95%CI) | 0.51 (0.34 – 0.77) | 1.00 (0.72 – 1.39) | 1.00 (Referent) | 1.29 (0.94 – 1.76) | 1.67 (1.24 – 2.25) |
| Lung – Female |  |  |  |  |  |
| Number of cases | 22 | 33 | 39 | 35 | 39 |
| HR (95%CI) | 0.55 (0.32 – 0.92) | 0.81 (0.51 – 1.28) | 1.00 (Referent) | 0.91 (0.58 – 1.44) | 1.00 (0.64 – 1.56) |
| Lung – Male |  |  |  |  |  |
| Number of cases | 56 | 64 | 79 | 75 | 86 |
| HR (95%CI) | 0.69 (0.49 – 0.97) | 0.80 (0.57 – 1.11) | 1.00 (Referent) | 1.01 (0.74 – 1.38) | 1.07 (0.79 – 1.45) |

Every SD increase in PRS is associated with 35-73% elevated risks of breast, prostate and colorectal cancers (P<2.19 × 10^−7^, **Table 3**). The increased risk for female and male lung cancer was lower than the other three cancers (HR_female_: 1.17 [1.01 to 1.36], p=4.07 × 10^−2^; HR_male_: 1.17 [1.06 to 1.29], p=1.52 × 10^−3^). Age at recruitment is significantly associated with elevated risks of developing all cancers, with the exception of female breast cancer (HR: 1.00 [0.99 to 1.02], p=0.571). Highest education level and BMI were positively correlated with breast cancer risk. Smoking was significantly associated with a ∼30% reduction in risk of prostate cancer, but increased the risk of lung cancer by approximately two- and five-fold for past and current smokers, compared to non-smokers, respectively. Alcohol consumption increased the risk of both female and male colorectal cancer by approximately 60% but was only significant for male colorectal cancer. Family history of cancer was only significantly associated with an increased risk for prostate cancer (HR: 1.61 [1.22 to 2.13], p=7.59 ×10^−4^).

**Table 3.** Associations between per standard deviation (SD) increase in site-specific polygenic risk scores and cancer occurrence. Hazard ratios (HR) and corresponding 95% confidence intervals (CI) were estimated using Cox proportional hazard models, adjusted for age at recruitment, dialect group, highest education attained, body mass index, smoking status, alcohol consumption, and physical activity. Follow-up time was censored at 20 years after recruitment. Significant results are shown in bold.

|  | Cancer site |  |  |  |  |  |  |  |  |  |  |  |
| --- | --- | --- | --- | --- | --- | --- | --- | --- | --- | --- | --- | --- |
|  | Breast |  | Prostate |  | Colorectal - female |  | Colorectal - male |  | Lung - female |  | Lung - male |  |
|  | HR (95% CI) | P-value | HR (95% CI) | P-value | HR (95% CI) | P-value | HR (95% CI) | P-value | HR (95% CI) | P-value | HR (95% CI) | P-value |
| Site-specific polygenic risk score, per SD increase | **1.46 (1.33 – 1.59)** | **3.83E-16** | **1.73 (1.54 – 1.95)** | **2.31E-20** | **1.35 (1.20 – 1.51)** | **2.19E-07** | **1.44 (1.30 – 1.59)** | **5.41E-12** | **1.17 (1.01 – 1.36)** | **4.07E-02** | **1.17 (1.06 – 1.29)** | **2.52E-03** |
| Age at recruitment, years | 1.00 (0.99 – 1.02) | 5.71E-01 | **1.08 (1.06 – 1.10)** | **9.37E-22** | **1.07 (1.05 – 1.09)** | **1.00E-16** | **1.06 (1.05 – 1.08)** | **9.53E-18** | **1.07 (1.05 – 1.10)** | **1.90E-10** | **1.09 (1.07 – 1.10)** | **2.54E-27** |
| Dialect group (Cantonese vs Hokkien) | 0.90 (0.74 – 1.08) | 2.47E-01 | 0.98 (0.77 – 1.23) | 8.35E-01 | 0.80 (0.64 – 1.01) | 6.47E-02 | 1.22 (0.99 – 1.50) | 6.78E-02 | 0.92 (0.67 – 1.26) | 5.97E-01 | 1.06 (0.86 – 1.31) | 5.95E-01 |
| Highest education (Primary vs No) | 1.21 (0.95 – 1.53) | 1.25E-01 | 1.28 (0.79 – 2.08) | 3.19E-01 | 1.08 (0.83 – 1.40) | 5.73E-01 | 0.98 (0.70 – 1.37) | 8.91E-01 | 0.83 (0.58 – 1.19) | 3.14E-01 | 0.90 (0.66 – 1.22) | 4.98E-01 |
| Highest education (Secondary or above vs No) | **1.54 (1.18 – 2.01)** | **1.55E-03** | 1.54 (0.93 – 2.53) | 9.01E-02 | 1.06 (0.76 – 1.48) | 7.41E-01 | 0.80 (0.55 – 1.16) | 2.33E-01 | 1.09 (0.69 – 1.73) | 7.18E-01 | 0.65 (0.46 – 0.93) | 1.89E-02 |
| Body mass index, kg/m<sup>2</sup> | **1.04 (1.02 – 1.07)** | **9.12E-04** | 1.01 (0.98 – 1.05) | 4.51E-01 | 0.99 (0.96 – 1.02) | 5.42E-01 | 1.02 (0.98 – 1.05) | 3.19E-01 | 0.97 (0.92 – 1.01) | 1.57E-01 | 0.96 (0.93 – 1.00) | 4.93E-02 |
| Smoking status (Ex-smoker vs Non-smoker) | 0.90 (0.44 – 1.82) | 7.70E-01 | **0.71 (0.52 – 0.96)** | **2.42E-02** | 1.50 (0.85 – 2.64) | 1.59E-01 | 1.17 (0.90 – 1.52) | 2.36E-01 | **2.16 (1.04 – 4.48)** | **3.82E-02** | **1.98 (1.39 – 2.80)** | **1.32E-04** |
| Smoking status (Current smoker vs Non-smoker) | 0.84 (0.50 – 1.41) | 4.99E-01 | **0.72 (0.54 – 0.97)** | **2.85E-02** | 1.12 (0.70 – 1.78) | 6.36E-01 | 1.22 (0.96 – 1.56) | 1.08E-01 | **5.71 (3.93 – 8.29)** | **4.81E-20** | **5.02 (3.74 – 6.74)** | **6.26E-27** |
| Alcohol consumption (Weekly vs Never/ Occasionally) | 1.04 (0.65 – 1.67) | 8.68E-01 | 0.96 (0.68 – 1.36) | 8.31E-01 | 0.76 (0.38 – 1.54) | 4.45E-01 | 1.31 (1.00 – 1.73) | 5.39E-02 | 0.72 (0.27 – 1.94) | 5.12E-01 | 0.90 (0.66 – 1.23) | 5.18E-01 |
| Alcohol consumption (Daily vs Never/ Occasionally) | 0.73 (0.27 – 1.97) | 5.39E-01 | 0.70 (0.38 – 1.29) | 2.54E-01 | 1.60 (0.71 – 3.60) | 2.58E-01 | **1.64 (1.15 – 2.34)** | **6.54E-03** | 0.67 (0.17 – 2.72) | 5.76E-01 | 1.23 (0.87 – 1.76) | 2.42E-01 |
| Moderate physical activity (1-3 hours/week vs No) | 0.98 (0.75 – 1.27) | 8.61E-01 | 1.16 (0.87 – 1.56) | 3.13E-01 | 0.89 (0.63 – 1.24) | 4.81E-01 | 1.02 (0.78 – 1.35) | 8.79E-01 | 1.01 (0.63 – 1.61) | 9.62E-01 | 0.88 (0.65 – 1.20) | 4.32E-01 |
| Moderate physical activity (≥3 hours/week vs No) | 1.18 (0.86 – 1.61) | 3.03E-01 | 1.05 (0.72 – 1.54) | 7.81E-01 | **0.60 (0.37 – 0.97)** | **3.57E-02** | 1.10 (0.80 – 1.52) | 5.45E-01 | 0.95 (0.53 – 1.68) | 8.52E-01 | 0.84 (0.57 – 1.22) | 3.51E-01 |
| Vigorous physical activity/ strenuous sports at least once a week (Yes vs No) | 1.24 (0.90 – 1.70) | 1.88E-01 | 1.09 (0.82 – 1.45) | 5.65E-01 | 1.08 (0.67 – 1.72) | 7.62E-01 | 0.75 (0.57 – 1.00) | 5.06E-02 | 0.57 (0.23 – 1.40) | 2.22E-01 | 0.94 (0.71 – 1.25) | 6.77E-01 |
| Family history (Yes vs No) | 1.15 (0.91 – 1.45) | 2.53E-01 | **1.61 (1.22 – 2.13)** | **7.59E-04** | 1.08 (0.79 – 1.48) | 6.17E-01 | 1.24 (0.95 – 1.62) | 1.09E-01 | 0.67 (0.40 – 1.13) | 1.36E-01 | 0.96 (0.71 – 1.32) | 8.15E-01 |

### Number of cancers that developed within PRS at-risk groups

Modelling (Cox proportional hazards) the risk of developing cancer using standardized PRS and accounting for age at recruitment, 14-23% of participants were at a greater than 1.5 risk of developing prostate (23%), female breast (14%) and male colorectal cancer (14%) (**Table 4**). The proportions were lower for female colorectal (6%) and lung cancer (1%). The number of participants who developed site-specific cancers in the at-risk group represented 42%, 25%, 22%, 11%, and 1% for prostate, female breast, male colorectal, female colorectal, and lung cancers, respectively. Among 1,674 women who were associated with HR>1.5 based on per standard deviation increase of PRS, 115 breast cancers (6.9%) developed during the follow-up. This proportion is nearly twice that of women not identified to be at high risk (380/10,410, 3.7%). Among 2,220 men who were associated with HR>1.5 based on per standard deviation increase of PRS, 120 prostate cancers (5.4%) developed during the follow-up. This proportion is over twice that of men not identified to be at high risk (118/7,390, 2.5%).

**Table 4.** Number of individuals estimated to have a hazard ratio (HR) associated with per standard deviation increase in site-specific polygenic risk score above the arbitrary threshold (1.5, 2.0, 2.5 and 3.0). To estimate the HR for each individual, we applied the *predict()* function with option *type=“risk”* to the Cox model with PRS (standardised to mean 0 and variance 1) and age at recruitment.

| Cancer site - gender | HR ≥ 1.5 |  | HR ≥ 2.0 |  | HR ≥ 2.5 |  | HR ≥ 3.0 |  |
| --- | --- | --- | --- | --- | --- | --- | --- | --- |
|  | *n*<br>(% of study<br>population) | Number who<br>developed cancer<br>(% of total cases) | *n*<br>(% of study<br>population) | Number who<br>developed cancer<br>(% of total cases) | *n*<br>(% of total in<br>SCHS) | Number who<br>developed cancer<br>(% of total cases) | *n*<br>(% of total in<br>SCHS) | Number who<br>developed cancer<br>(% of total cases) |
| Effect of PRS alone |  |  |  |  |  |  |  |  |
| Breast – female | 1674 (14) | 115 (25) | 373 (3) | 27 (6) | 84 (1) | 10 (2) | 27 (0) | 2 (0) |
| Prostate – male | 2220 (23) | 120 (42) | 981 (10) | 61 (21) | 439 (5) | 32 (11) | 210 (2) | 19 (7) |
| Colorectal – female | 756 (6) | 33 (11) | 48 (0) | 0 (0) | 1 (0) | 0 (0) | 0 (0) | 0 (0) |
| Colorectal – male | 1300 (14) | 82 (22) | 291 (3) | 23 (6) | 72 (1) | 10 (3) | 15 (0) | 3 (1) |
| Lung – female | 73 (1) | 1 (1) | 0 (0) | 0 (0) | 0 (0) | 0 (0) | 0 (0) | 0 (0) |
| Lung – male | 52 (1) | 2 (1) | 0 (0) | 0 (0) | 0 (0) | 0 (0) | 0 (0) | 0 (0) |
| Total – females | 2326 (19) | 148 (19) | 418 (3) | 27 (4) | 84 (1) | 10 (1) | 27 (0) | 2 (0) |
| Total – males | 3250 (34) | 204 (20) | 1240 (13) | 84 (8) | 509 (5) | 42 (4) | 224 (2) | 22 (2) |
| Adjusted for age at recruitment |  |  |  |  |  |  |  |  |
| Breast – female | 1698 (14) | 112 (24) | 373 (3) | 26 (6) | 86 (1) | 10 (2) | 29 (0) | 2 (0) |
| Prostate – male | 2854 (30) | 158 (55) | 1834 (19) | 118 (41) | 1192 (12) | 83 (29) | 817 (9) | 63 (22) |
| Colorectal – female | 2887 (24) | 138 (45) | 1561 (13) | 92 (30) | 850 (7) | 57 (18) | 447 (4) | 34 (11) |
| Colorectal – male | 2510 (26) | 175 (46) | 1443 (15) | 119 (31) | 832 (9) | 71 (19) | 476 (5) | 39 (10) |
| Lung – female | 3279 (27) | 91 (54) | 2104 (17) | 62 (37) | 1360 (11) | 42 (25) | 925 (8) | 29 (17) |
| Lung – male | 2732 (28) | 177 (49) | 1687 (18) | 107 (30) | 1035 (11) | 73 (20) | 653 (7) | 41 (11) |
| Total – females | 4273 (35) | 249 (33) | 1903 (16) | 118 (15) | 933 (8) | 67 (9) | 476 (4) | 36 (5) |
| Total – males | 4189 (44) | 498 (50) | 2821 (29) | 336 (34) | 1914 (20) | 221 (22) | 1324 (14) | 140 (14) |
| Fully adjusted for all covariates |  |  |  |  |  |  |  |  |
| Breast – female | 2209 (18) | 150 (32) | 760 (6) | 61 (13) | 257 (2) | 25 (5) | 86 (1) | 12 (3) |
| Prostate – male | 3000 (31) | 179 (62) | 1947 (20) | 135 (47) | 1319 (14) | 105 (36) | 956 (10) | 75 (26) |
| Colorectal – female | 3006 (25) | 143 (46) | 1663 (14) | 94 (30) | 935 (8) | 60 (19) | 548 (5) | 37 (12) |
| Colorectal – male | 2803 (29) | 204 (54) | 1666 (17) | 134 (35) | 1041 (11) | 97 (26) | 640 (7) | 58 (15) |
| Lung – female | 3111 (26) | 98 (58) | 2027 (17) | 78 (46) | 1402 (12) | 64 (38) | 1062 (9) | 59 (35) |
| Lung – male | 3572 (37) | 265 (74) | 2734 (28) | 232 (64) | 2134 (22) | 206 (57) | 1684 (18) | 180 (50) |
| Total – females | 4783 (40) | 291 (38) | 2363 (20) | 155 (20) | 1181 (10) | 85 (11) | 633 (5) | 49 (6) |
| Total – males | 5542 (58) | 633 (63) | 4191 (44) | 492 (49) | 3247 (34) | 400 (40) | 2547 (27) | 307 (31) |

When age at recruitment was included in the models, 14-44% of the participants were at a greater than 1.5 risk of developing the various cancers. The number of participants who developed site-specific cancers in the at-risk group increased to 24-55%. In the fully adjusted models, 18-58% the participants were at a greater than 1.5 risk of developing the various cancers. The number of participants who developed site-specific cancers in the at-risk group increased further to 32-74%. All Cox models presented in **Tables 2, 3** and **4** did not violate the proportionality assumption for the PRS studied (p-values of *cox-zph()* for PRS were >0.05).

### PRS discriminatory ability

The highest AUC obtained from logistic models was observed for prostate cancer (0.66, 95% CI: [0.62 to 0.69]), followed by female breast cancer (0.61 [0.58 to 0.63]), male colorectal cancer (0.60, 95% CI = 0.58 to 0.63), female colorectal cancer (0.58 [0.54 to 0.61]), male lung cancer (0.55 [0.52 to 0.58]) and female lung cancer (0.55 [0.50 to 0.59]) (**Figure 1B**).

### PRS predictive ability

In terms of the five-year absolute risk of developing site-specific cancers, the largest difference between the highest and lowest PRS categories was observed for prostate cancer, followed by breast cancer (**Figure 1C**). A separation of the absolute risk curves was observed for female breast cancer already at age 30 years. For prostate cancer, the separation of curves was observed only after age 50 years. Slight separation of the curves began after 50 years of age for colorectal and lung cancer.

### PRS calibration

In general, predicted risks for the higher PRS categories did not correspond well to the observed proportions for female breast, prostate, and female lung cancers (**Figure 1D**); in particular, predicted risks were overestimated for the higher risk categories. Overestimation of risk was observed for all PRS categories for male lung cancer. In contrast, predicted risks were underestimated for both female and male colorectal cancers.

## DISCUSSION

Precision prevention in oncology is based on the idea that an individual’s risk, which is influenced by genetics, environment, and lifestyle factors, is linked to the amount of benefit achieved through cancer screening [28]. Risk stratification for cancer screening can be used in this framework to identify and recommend screening for persons with a high enough cancer risk that the benefits outweigh the risks. Several PRS prediction models have been established for site-specific cancers, each with its own set of strengths and limitations, and different risk models may produce different results for the same individual.

In an increasingly inclusive world, genetic studies fall short on diversity. According to a 2009 study, an overwhelming 96% of people who took part in genome-wide association studies (GWAS) were of European ancestry [29]. GWAS results are the backbone on which PRS is developed. A concern raised was that, without representation from a broader spectrum of populations, genomic medicine may be limited to benefitting “a privileged few” [30].

Genetic studies in 2016 showed that the proportion of people not of European ancestry included in GWAS has increased to approximately 20% [30]. Most of this rise can be attributed to more research on Asian ancestry communities in Asia [30]. With increasing interest worldwide in using a risk-based approach to screening programs over the current age-based paradigm, this progress raises questions on whether selected established PRS shown to perform well in European-based populations has equal utility in Asians. Nonetheless, as our results show, most of the populations from which PRS were developed are still predominantly of European ancestry.

In accordance with published Polygenic Risk Score Reporting Standards, we reported PRS distribution, discrimination, predictive ability, and calibration for each of the four common cancers studied [31]. Our results show that cancer cases were associated with higher PRS compared to non-cancer controls. In the age-adjusted models, a constant trend between PRS percentile rank and observed cancer risk in our study population supports the validity of PRS for breast, prostate, and colorectal cancers, but not for lung cancer. The best-performing PRS for female breast cancer was able to stratify women into distinct bands of breast cancer risk at an earlier age, and across all ages, suggesting that it could be a useful prediction tool in risk-based breast cancer screening in combination with other risk factors specific to breast cancer [17]. This PRS has been incorporated into a pilot risk-based breast cancer screening study in a comparable study population [32]. The best performing PRS for prostate and male colorectal cancers in this study appeared to exhibit sufficient discriminatory ability and predictive value, especially for older participants.

PRS may be of limited use in predicting female colorectal and female/male lung cancer. The least predictive value was in lung cancer, which could be related to the higher prevalence of EGFR mutant lung cancer which has an Asian predilection, thus less amenable to PRS developed in Caucasian population [33]. For these patients <10% of population were identified with >1.5 HR of developing incident cancers.

There is room for improvement in the discriminatory ability of PRS [34]. As noted by Lambert et al in a review, a wider divergence between the average scores of cases and non-cases (quantified by AUC) and associated effect sizes (odds ratio and standard deviation) is expected when PRS explains more of the heredity for each trait [2]. Larger GWAS sample sizes of appropriate ancestries and the inclusion of rarer genetic variants, obtained through other methods such as whole-genome sequencing, would likely be required to boost explained heritability [2]. In addition, group-wise estimates, which arbitrarily classify the top 10%, 5%, or 1% of samples as the at-risk group, are not optimal for decisions at the individual level [34]. Emerging new methodologies that estimate probability values for hypothetically assigning an individual as at risk or not at risk, thus providing individuals with more clarity, may help to overcome this limitation [35]. At this point, PRS may not have yet reached the standards as a clinical tool by itself. However, it is still helpful in guiding screening decisions and supplementing established protocols [1].

As highlighted by Wei et al, the reliability of score values is necessary for application at the individual level [36]. Even when the PRS have adequate discrimination, estimated risks can be unreliable [37]. Our results show that cancer risk estimates based on PRS developed using populations of European ancestry are not optimally calibrated for our Asian study population. Poorly calibrated PRS can be misleading and have clinical repercussions [37, 38]. Underestimation of risk may result in a false sense of security. Overestimation of risk may cause unnecessary anxiety, misguided interventions, and overtreatment. In a population-wide screening setting, however, where the return of PRS results can be designed such that only high-risk individuals are highlighted, underestimation of risk may be less of an issue. Arguably, with parallel input from other risk factors and evaluation by healthcare specialists, the overestimation of risk that results in a higher number of at-risk individuals identified may increase the number of cancers potentially detected early. Nonetheless, suitable correction factors will be required to ensure the reliability of PRS prior to clinical implementation.

While the study population used in this analysis comprises less than a thousand cases of the most common cancers examined, the Singapore Chinese Health Study, established between April 1993 and December 1998, is one of the largest population-based Asian cohorts in the world with high-quality prospective data on exposure and comprehensive capture of morbidity and mortality. All cancer cases are incident cases diagnosed over three decades of follow-up. This is one of the best resources to evaluate the utility of PRS in a prospective manner. The findings open a window in our current understanding of which PRS is relevant and ready to be deployed in risk-based cancer screening studies.

Ethnic representation in PRS model development, PRS validation, limited discriminative ability in the general population, ill calibration, insufficient healthcare professional and patient education, and healthcare system integration are all hurdles that must be crossed before PRS can be implemented responsibly as a public health instrument [39, 40]. Importantly, genetic literacy will be a critical prerequisite for the successful implementation of PRS in population-based health screening. It is pivotal that uncertainty associated with risk estimates derived from PRS is communicated clearly [1]. In addition, an individual flagged to be at high risk of developing cancer may be unaware of the range of surveillance options available [41]. In a commentary evaluating the “right not to know” in genomics research by Gold and Green, it was noted that among those who chose not to have their results returned, nearly half of them changed their minds after an education intervention [42].

While nationwide screening programs have helped to raise cancer awareness, there is still a need to improve the effectiveness and efficiency of cancer screening in Asian countries such as Singapore, given the steadily rising incidence rates. Despite the challenges, a risk-based screening strategy that includes the use of PRS should be actively examined for research and implementation.

## Data Availability

All data produced in the present study are available upon reasonable request to the authors

## DECLARATIONS

### Ethics approval and consent to participate

The study was approved by the institutional review boards of the University of Southern California, the National University of Singapore, and the Agency for Science, Technology and Research (A*STAR, reference number 2022-042). Written, informed consent was obtained from all study participants.

### Consent for publication

Not applicable.

### Availability of data and materials

All polygenic risk scores used in this study are publicly available in the PGS Catalog (https://www.pgscatalog.org).

The data that support the findings of our study are available from the corresponding authors of the study upon reasonable request (Dr Rajkumar s/o Dorajoo, and Dr Jingmei Li). More information regarding the data access to SCHS can be found at: https://sph.nus.edu.sg/research/cohort-schs/. The data are not publicly available due to Singapore laws.

Source Data 1 contain the numerical data used to generate the figure 1.

The code for the study is uploaded as Source Code 1.

### Authors’ contributions

Conception or design of work: Jingmei Li, Peh Joo Ho, Rajkumar s/o Dorajoo

Acquisition of resources for the generation of data, identification of outcomes via linkage and supervision for the collection of data: Woon Puay Koh (PI of the Singapore Chinese Health Study) Data acquisition: Jingmei Li, Rajkumar s/o Dorajoo, Chiea Chuen Khor, Woon Puay Koh, Jian-Min Yuan

Interpretation of data: Jingmei Li, Peh Joo Ho, Rajkumar s/o Dorajoo, Iain Bee Huat Tan Drafting of manuscript: Jingmei Li, Peh Joo Ho, Rajkumar s/o Dorajoo

Manuscript approval: All authors

All authors agreed both to be personally accountable for the author’s own contributions and to ensure that questions related to the accuracy or integrity of any part of the work, even ones in which the author was not personally involved, are appropriately investigated, resolved, and the resolution documented in the literature.

## Acknowledgments

We thank the Singapore Cancer Registry for the identification of incident cancer cases among participants of the Singapore Chinese Health Study and Siew-Hong Low of the National University of Singapore for supervising the fieldwork of the Singapore Chinese Health Study.

## ADDITIONAL FILES

Additional File 1 - Supplementary tables.xlsx

